# COVID-19 antibody detection and assay performance using red cell agglutination

**DOI:** 10.1101/2021.06.09.21258232

**Authors:** Kshitij Srivastava, Kamille A West, Valeria De Giorgi, Michael R Holbrook, Nicolai V. Bovin, Stephen M Henry, Willy A Flegel

## Abstract

Red cells can be labelled with peptides from the SARS-CoV-2 spike protein and used for serologic screening of SARS-CoV-2 antibodies. We evaluated 140 convalescent COVID-19 patients and 275 healthy controls using this C19-kodecyte assay. The analytical performance of the new assay was compared with a virus neutralizing assay and 2 commercial chemiluminescent antibody tests (Total assay and IgG assay, Ortho). The C19-kodecyte assay detected SARS-CoV-2 antibodies with a sensitivity of 92.8% and specificity of 96.3%, well within the minimum performance range required by FDA for EUA authorization of serologic tests. The Cohen’s kappa coefficient was 0.90 indicating an almost perfect agreement with the Total assay. The Pearson correlation coefficient was 0.20 with the neutralizing assay (0.49 with IgG, and 0.41 with Total assays). The limited correlation in assay reaction strengths suggested that the assays may detect different antibody specificities. Our easily scalable C19-kodecyte assay may vastly improve test capacity in blood typing laboratories using their routine setups for column agglutination technique.

## Background

The coronavirus disease 2019 (COVID-19) pandemic, caused by the novel betacoronavirus SARS-CoV-2, has resulted in global public health crisis. As of May 27 2021, there have been more than 168 million confirmed cases of COVID-19, including more than 3 million deaths reported globally [1]. Vaccines against SARS-CoV-2 were first approved in China on June 25, 2020 for limited use in the military [2], followed by Russia on August 11, 2020 for emergency use [3]. In the United States, the United States Food and Drug Administration (FDA) issued the first emergency use authorization (EUA) for Pfizer-BioNTech COVID-19 vaccine on December 11, 2020 [4]. Worldwide, 15 COVID-19 vaccines have been authorized or approved, as of May 24, 2021 [5].

On August 23, 2020, the FDA issued an EUA for COVID-19 convalescent plasma (CCP) for the treatment of hospitalized COVID-19 patients [6]. CCP provides passive immunity through neutralizing antibodies against SARS-CoV-2 virus, helps control infection and modulates inflammatory response [7]. Only high-titer CCP to hospitalized older COVID-19 patients early in the disease course has been shown to reduce the progression of disease [6, 8]. However, recent studies have concluded no clear benefit to the CCP therapy [9] with many blood centers suspending collection of convalescent plasma.

Serologic assays detecting antibodies against the SARS-CoV-2 spike or nucleocapsid proteins have been developed to advance our understanding of the prognosis and clinical course of the disease, contribute to epidemiological research and aid in vaccine development. These serologic assays are also used to identify asymptomatic individuals, evaluate immune response in patients, identify of high-titer CCP units, determine duration and magnitude of immunity conferred by SARS-CoV-2 vaccine, and assist in the prediction of disease progression and epidemiology [10, 11]. The FDA has approved 79 SARS-CoV-2 antibody assays under EUA [12]. Only 11 of these serologic assays were accepted in the manufacture of high-titer COVID-19 convalescent plasma [6].

We recently developed a red cell agglutination-based assay to detect SARS-CoV-2 antibodies [13]. The assay uses peptide fragments of the SARS-CoV-2 spike protein to label red cells (C19-kodecytes). These C19-kodecytes are agglutinated in the presence of SARS-CoV-2 antibodies using routine serologic platforms. We perform a clinical evaluation of this C19-kodecyte assay in CCP donors previously assessed with 2 commercial immunoassays and a virus neutralizing assay [14].

## Materials and Methods

### Study population

COVID-19 convalescent plasma or EDTA anticoagulated whole blood was prospectively collected between April 2020 and January 2021 from 140 donors (2 asymptomatic and 138 with mild to severe disease) [14], who had previously tested positive for SARS-CoV-2 infection by real-time reverse transcription polymerase chain reaction (rRT-PCR). Plasma samples were collected from 125 consecutive Ortho Total negative healthy volunteer donors. An additional 150 plasma samples, collected in 2008, more than a decade before the COVID-19 outbreak, were also included as negative controls to evaluate specificity of the C19-kodecyte assay. An individual vaccinated with Moderna COVID-19 vaccine was prospectively analyzed for antibody response using the C19-kodecyte assay. Plasma, erythrocytes, and buffy coat aliquots were separated and stored at -80 °C (plasma) or in the vapor phase of liquid nitrogen containers (−150 °C; erythrocytes and buffy coat).

Institutional Review Board (IRB) approved protocols were NCT04360278, NCT00001846 and NCT00067054, respectively, which entailed written informed consent. The plasma controls from 2008 were collected under the Vaccine Research Center’s (VRC), National Institutes of Allergy and Infectious Diseases (NIAID), National Institutes of Health sample collection protocol VRC 200 (NCT00067054) in compliance with the NIH IRB approved procedures. All subjects met protocol eligibility criteria and agreed to participate in the study by signing the NIH IRB approved informed consent. Research studies with these samples were conducted by protecting the rights and privacy of the study participants.

### Anti-SARS-CoV-2 assays

Two chemiluminescent immunoassays directed against the SARS-CoV-2 spike glycoprotein, Ortho IgG (80.8% overall sensitivity and 98.1% overall specificity) [15], and Ortho Total (IgG, IgA and IgM; 87.7% overall sensitivity and 95.2% overall specificity) [15] (Ortho Clinical Diagnostics, Raritan, NJ, USA), authorized by FDA under an EUA for use by laboratories certified under Clinical Laboratory Improvement Amendments of 1988 (CLIA), were used to detect total binding antibodies against the SARS-CoV-2 virus in all 140 CCP donors [14, 15]. The 125 healthy volunteer donors were only tested with the Ortho Total assay. The 150 pre-COVID-19 plasma samples were presumed negative for SARS-CoV-2 and not tested with either of the immunoassays.

### SARS-CoV-2 virus neutralizing assay

The surface-exposed location of the spike glycoprotein makes it a major target of neutralizing antibodies [16]. An in-house developed fluorescence reduction neutralizing assay (FRNA; not authorized by FDA; sensitivity and specificity data not available) [17] was performed to detect neutralizing antibodies against the SARS-CoV-2 virus (NT50) in all 140 CCP samples [14].

### SARS-CoV-2 kodecytes

Dual epitope bearing kodecytes were prepared using a blend of FSL (Function-Spacer Lipid) constructs 1147 and 1255 (peptides fragments of the SARS-CoV-2 spike protein) at concentrations 1.5 μM and 2.5 μM, respectively (C19-kodecytes) [13]. FSL construct 808 (5 μM), with weaker reaction strength as compared to constructs 1147 and 1255 [13], was also tested in parallel. All samples reactive to C19-kodecytes were also tested against control cells, either unmodified red cells or kodecytes labelled with an unrelated FSL construct [13].

### Gel card column agglutination technique (CAT)

The RBC-antibody mixtures were prepared directly in the gel card wells (ID Cards LISS/Coombs, no. 50531; Bio-Rad Laboratories, Hercules, CA, USA) with 50 µL of SARS-CoV-2 kodecytes at 1% concentration and 25 µL of antibody solution. Gel cards were incubated at 37 °C for 15 min and centrifuged at 1,032 rpm for 10 minutes in an Ortho Workstation (Ortho Clinical Diagnostics, Raritan, NJ) at room temperature. All reactions were read macroscopically. A titer was determined as the highest dilution showing 1+* (weak positive) agglutination. The grading of the agglutination reaction was scored as 12, 10, 8, 5, 3, 0 for titer 4+, 3+, 2+, 1+, 1+* and negative, respectively [13, 18]. A higher score can indicate the presence of a higher antibody concentration. more antigens on the red cells, or a stronger antigen-antibody affinity. Total score was calculated by adding the individual scores at two-fold serial dilutions.

### Sensitivity and specificity

The sensitivity of our assay was evaluated on consecutive CCP samples collected from participants at various time points after rRT-PCR confirmation. Assay sensitivity was calculated as the percentage of samples that tested positive with the C19-kodecyte assay relative to the total number of PCR-confirmed COVID-19 positive samples [Sensitivity = True Positive / (True Positive + False Negative)].

The specificity of our assay was calculated as the percentage of samples that tested negative with the C19-kodecyte assay relative to the total number of COVID-19 negative samples [Specificity = True Negative / (True Negative + False Positive)].

### Statistical analyses

Unless stated otherwise, continuous data are given as median (Quartile 1 – Quartile 3). Confidence intervals (95% CI) were calculated according to the efficient-score method (corrected for continuity) [19]. Cohen’s kappa was used to evaluate between methods agreements [20, 21]. Receiver operating characteristic (ROC) curves were used to evaluate overall performance of the 2 Ortho and the C19-kodecyte assays.

## Results

A total of 140 CCP samples and 275 healthy controls were analysed in the C19-kodecyte assay (Supplementary Table S1). The median time interval from the onset of symptoms and plasma donation was 78 days (Quartile 1 – Quartile 3, 53 – 111 days; range, 33 – 331 days). We compared our results with those of Ortho chemiluminescent immunoassays and with a virus neutralizing assay.

### Ortho SARS-CoV-2 assays

Ortho IgG assay was positive in 92.1% of CCP samples (129/140) and Ortho Total assay was positive in 97.8% of CCP samples (137/140). The median log anti-SARS-CoV-2 Signal/Cut-off (S/CO) ratio and inter-quartile range for each assay were as follows: Ortho IgG (1.03, 0.73 – 1.22) and Ortho Total (2.46, 2.04 – 2.74) (Fig. 1).

**Fig. 1.**
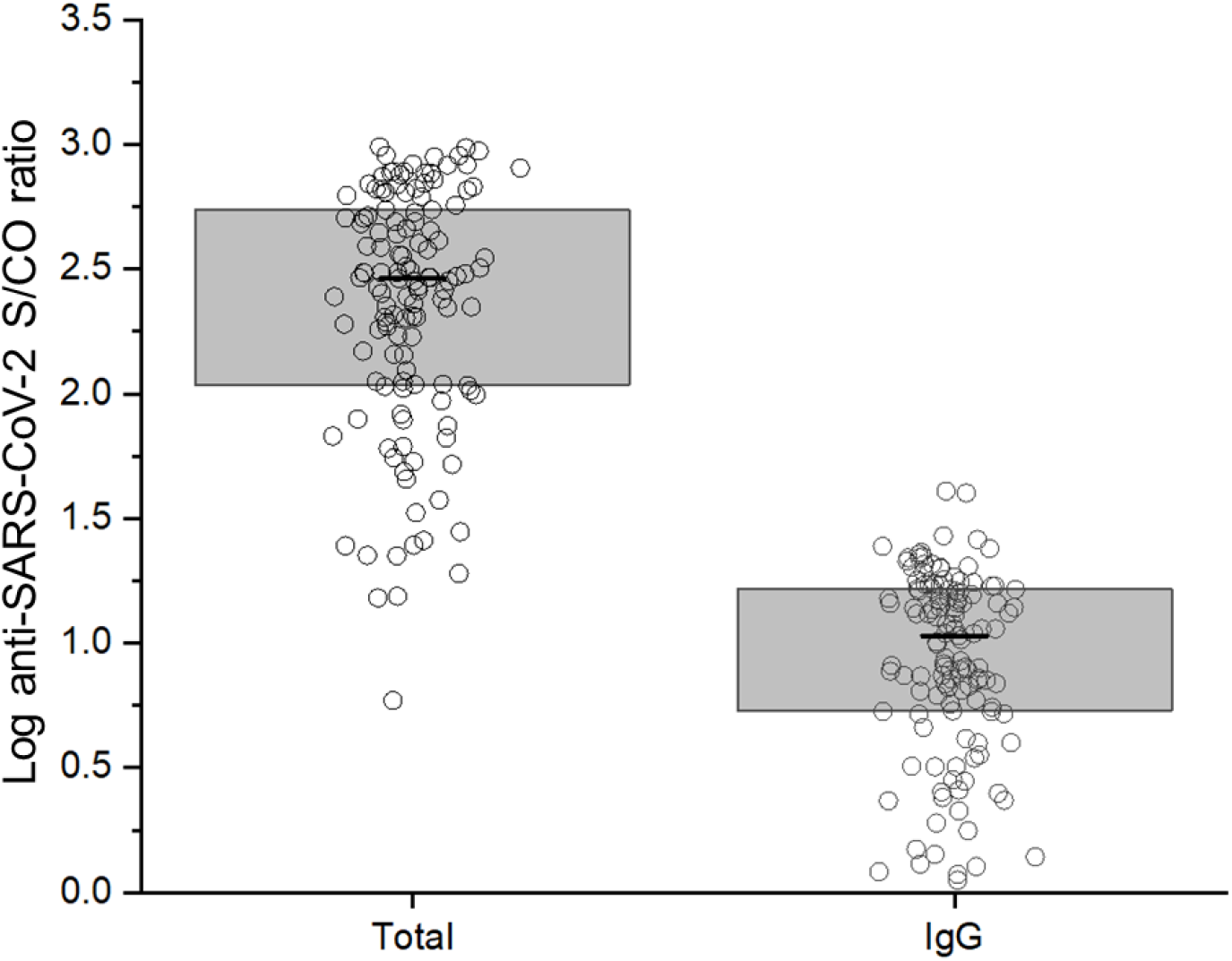
Anti-SARS-CoV-2 plasma S/CO ratios as measured using the Ortho Total and Ortho IgG assays. Shown are the antibody S/CO ratios against SARS-CoV-2 spike (S) protein in the convalescent plasma using the 2 Ortho assays. The horizontal bars indicate medians, and the shaded gray areas interquartile ranges. Each circle represents one patient.

### SARS-CoV-2 virus neutralizing assay

Of the 140 CCP samples tested, neutralizing data was not available for 40 (28.57%) samples. For the remaining 100 samples, there was a broad range of neutralization against live SARS-CoV-2, with 2 (1.43%) samples demonstrating high NT50 values of more than 600, and 5 (3.57%) samples with undetectable NT50 values of less than 40 (Supplementary Fig. S1).

### C19-kodecyte assay

Of the 140 CCP samples tested, C19-kodecyte assay was negative in 10 (7.1%) samples (Supplementary Table S3). The median log anti-SARS-CoV-2 score and inter-quartile range for C19-kodecyte assay were 1.41, 1.20 – 1.60, respectively. ROC curves were generated based on positive (140 SARS-CoV-2 rRT-PCR-positive) and negative results (125 Ortho Total negative and 150 pre-COVID-19 samples). The results showed that the area under the ROC curve (AUC) of C19-kodecyte reached 0.95 (95% CI: 0.93 -0.97) with sensitivity and specificity values of 92.8% (95% CI: 86.9% -96.3%) and 96.3% (95% CI: 93.2% -98.1%), respectively (Supplementary Fig. S2).

C19-kodecyte assay was used to test the antibody response in a vaccinated individual. The Ortho Total assay gave positive results 14 days after the 1^st^ vaccination and decreased 91 days later (data not shown). Using the C19-kodecyte assay, the individual’s plasma sample tested positive 28 days after the 1^st^ vaccination and increased a week later (score 40). The antibody concentration decreased when tested 91 days later (data not shown).

### Assay comparison

Almost all the CCP donors with longitudinal follow-up data (n = 40) showed a decrease in the antibody concentration from day 1 to their most recent donation, which varies from 7 to 44 weeks (Fig. 2). We compared the neutralizing activity in the CCP samples to C19-kodecyte score as well as 2 clinical diagnostic assays (Ortho Total and Ortho IgG). There was a universally positive relationship between NT50 and all 3 binding assays tested, with Spearman’s correlation r ranging from 0.20 to 0.49 and linear R^2^ values of 0.0796 to 0.2959 reflecting a weak linear relationship (Fig. 3). The strongest positive correlation between assays was the Ortho Total and Ortho IgG assays (Spearman’s r = 0.87, p = <0.001).

**Fig. 2.**
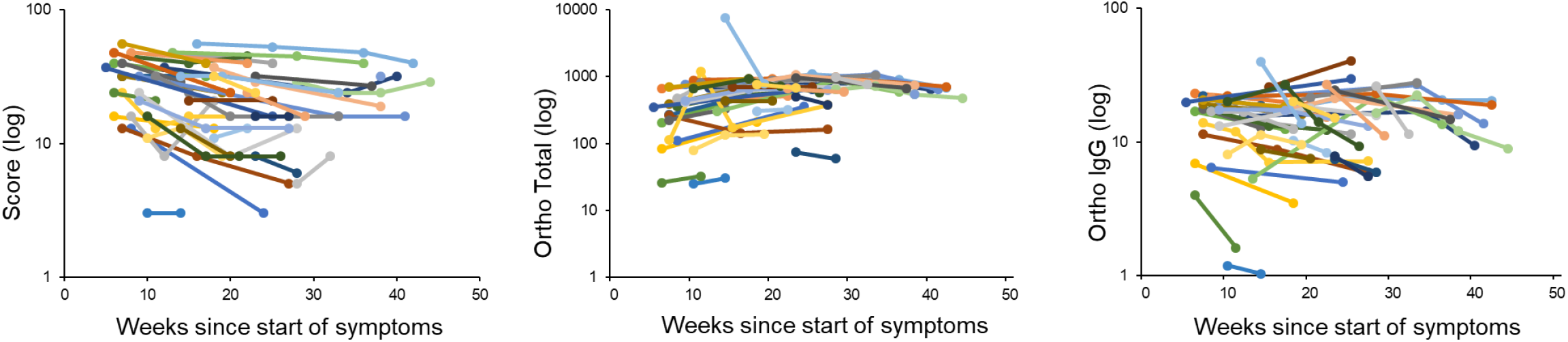
Correlation plots comparing all assays tested, with linear curve fitting R^2^ value shown. Spearman’s correlation (r) for pairwise comparisons of each assay tested is also shown. Each circle represents one patient. Total score was calculated by adding the individual scores at two-fold serial dilutions.

**Fig. 3.**
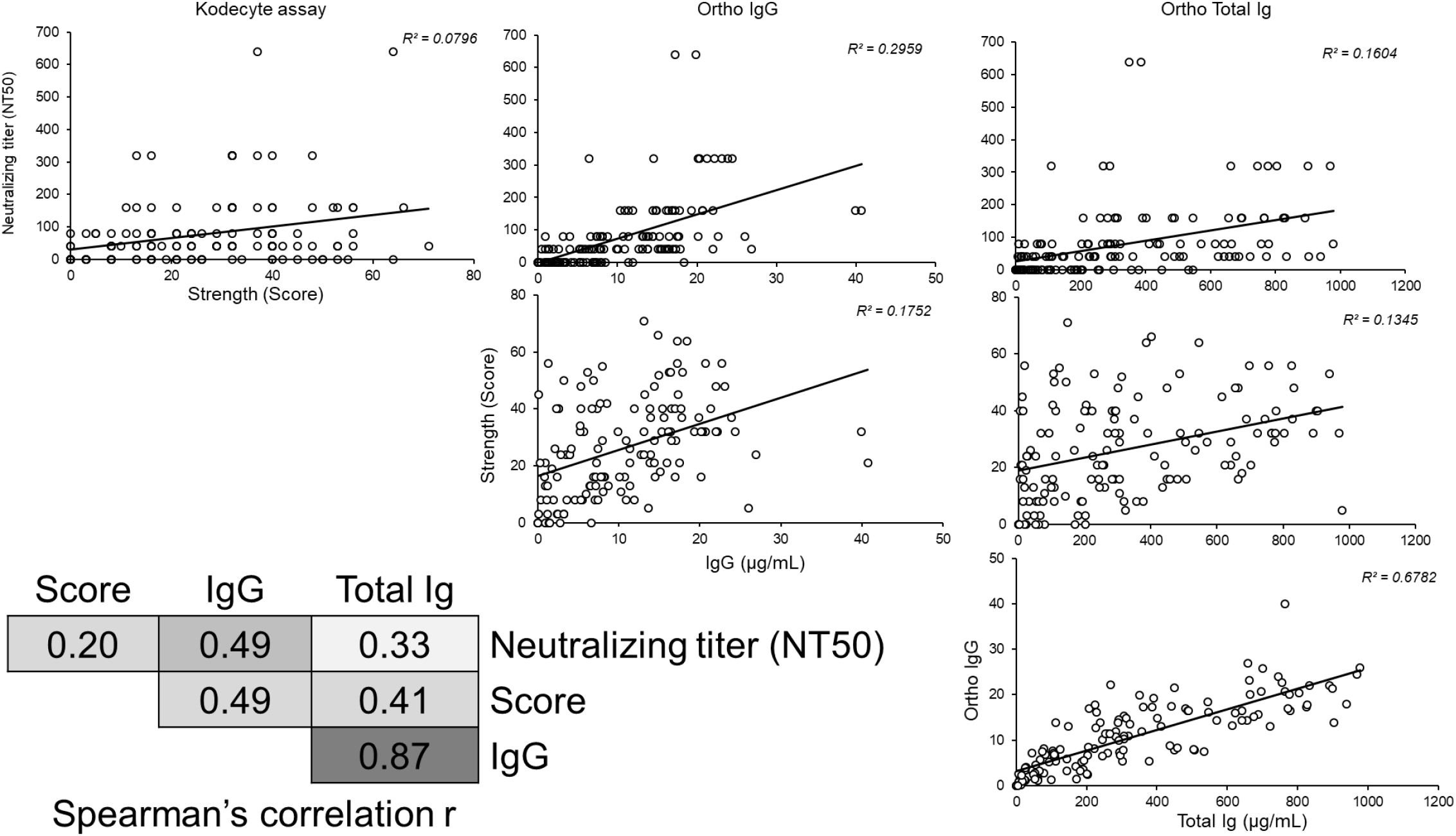
Change in Strength (score), Ortho Total and Ortho IgG S/CO ratio over time for each individual donor. Each data point represents the antibody levels calculated from an individual convalescent plasma donation, and each color represents an individual donor followed longitudinally. Each timepoint was calculated using the day of donation and the donor’s self-reported first day of symptoms. Total score was calculated by adding the individual scores at two-fold serial dilutions.

A few CCP samples were identified with discordant Ortho (Total and IgG) and neutralizing antibody S/CO ratios, including some with high neutralizing antibody and low Ortho antibody S/CO ratio in individual assays and vice versa (Supplementary Table S3). With positive and negative data available only for the C19-kodecyte and Ortho Total assays, the agreement between the 2 assays was considered as almost perfect [21], because the Cohen’s kappa was 0.90 (95% CI: 0.85-0.95) [20] (Supplementary Table S2).

There were 14 CCP donors that tested negative with at least one of the assays (Supplementary Table S3). We observed 3 CCP donors with no detectible antibody in the C19-kodecyte and both Ortho assays, without virus neutralizing assay data available. Additionally, C19-kodecyte assay was repeatedly positive in 10 healthy donors (6 Ortho Total negative and 4 pre-COVID-19).

## Discussion

The SARS-CoV-2 kodecyte based antibody diagnostic assay is a simple test for rapid screening of SARS-CoV-2 antibodies in human plasma [13]. The assay can be easily established in a blood bank, using its typical equipment, and is well suited for surveillance studies. We compared the C19-kodecyte assay with 2 established commercial assays (Ortho Total and Ortho IgG) and an in-house developed virus neutralization assay. All 4 assays detect antibodies directed against the SARS-CoV-2 spike glycoprotein [13, 15, 16].

In our study, the C19-kodecyte assay results demonstrated excellent analytical performance and exhibited almost perfect agreement with the Ortho Total assay (Cohen’s kappa = 0.90), albeit with limited correlation (0.41). However, a small proportion (14/140 CCP donors and 10/275 controls) of samples differed (Supplementary Table S3). This pattern may represent varied assay sensitivity, specificity, or a combination of both among the different assays as the C19-kodecyte assay primarily detects IgG antibody while the Ortho Total assay detects IgG, IgM and IgA antibodies. This may also suggest that COVID-19 patients develop a broad antibody repertoire against multiple SARS-CoV-2 proteins and epitopes and different assays detect diverse antibody specificities.

Another possible explanation for the weak correlation, despite excellent agreement, can be the difference in the underlying principle of the assays. The antigens (peptides) on the kodecytes are small, monospecific epitopes that react with a single antigen binding (Fab) region of antibody. Therefore, in contrast to recombinant protein diagnostic assays where multiple epitopes are present and are available to interact with different antibodies, C19-kodecytes with 2 epitopes (1147 and 1255), will probably only react with 2 very specific antibodies of the polyclonal antibody pool. Furthermore, it is speculated that kodecyte peptide epitopes are located in the glycocalyx of the red cell membrane and are surrounded by carbohydrates and proteins [13, 22], which may act like a natural buffer against low-affinity IgG antibodies and the lower affinity IgM antibodies and allows for the use of undiluted plasma samples.

A decrease in the SARS-CoV-2 antibody concentrations was shown for all CCP donors including the single vaccinated donor and has been reported previously in multiple studies [14, 23-26]. Antibody concentrations are also known to decay faster in individuals with mild infection or asymptomatic disease [27-29] resulting in no detectable SARS-CoV-2 antibodies in commercial antibody detection assays [30, 31].

The C19-kodecyte assay sensitivity (92.1%) was much greater than that published for the 2 Ortho assays (IgG: 80.8% and Total: 87.7%) [15]. The C19-kodecyte assay also demonstrated a high specificity (95.2%) and did not differ significantly from the Ortho assays (IgG: 98.1% and Total: 95.2%). The sensitivity and specificity values of the C19-kodecyte assay are well within the minimum performance range of FDA issued EUA authorized serology tests (90% sensitivity and 95% specificity) [32].

The C19-kodecyte assay, although a beta version with a possibility of further refinement, is comparable to other FDA authorized or approved assays. However, an optimization can still be attempted and improvements seem achievable for sensitivity and specificity by adjusting the sequence of peptides and the FSL construct concentrations. Slightly changing the existing peptide sequences by shifting or elongating them either left or right into the flanking sequences has the potential to improve both sensitivity and specificity [33]. Alternatively, different peptides could be evaluated and when testing CCP samples with a different SARS-CoV-2 spike protein sequence (Peptide 808), 3 samples that tested false negative in the C19-kodecyte assay tested positive with Peptide 808-kodecyte assay (Supplementary Table S3). Likewise, optimization of the concentration of peptides during kodecyte preparation can also improve reaction specificity of the assay.

The unusually flexibility of the C19-kodecyte assay can easily be adapted using different peptides from the SARS-CoV-2 spike and nucleocapsid proteins. This approach allows to modify the assay specificity to determine the duration and magnitude of immunity conferred by SARS-CoV-2 vaccine which would be an advantage for population surveillance for ongoing infection and maintenance of immunity.

In summary, we performed an analytical validation of the C19-kodecyte SARS-CoV-2 immunoassay in 140 clinical samples. Our easily scalable C19-kodecyte assay can be operated in blood typing laboratories worldwide using routine setups for column agglutination or tube techniques [13] to evaluate patients and convalescent donors. The technique could vastly improve assay capacity, particularly in countries with under-developed health systems.

## Data Availability

data as documented in the manuscript and supplement

## Acknowledgments

The authors thank Marina U Bueno for sample coordination and Sita Shrestha and Nadine R Dowling for technical support in the serologic testing. We acknowledge the contributions of our colleagues Martin R Gaudinski, Julie E Ledgerwood, Maria B Florez, Emily E Coates and Ingelise J Gordon at the Vaccine Research Center (VRC) Clinical Trials Program, NIAID and Robin A Gross, Janie Y Liang, Steven D Mazur and Elena N Postnikova at the Integrated Research Facility (IRF), Frederick, NIAID. This work was supported in whole or in part by the Intramural Research Program (project ID ZIC CL002128) of the NIH Clinical Center at the National Institutes of Health; National Institute of Allergy and Infectious Diseases, National Institutes of Health, U.S. Department of Health and Human Services (DHHS), under Contract No. HHSN272201800013C; and by the New Zealand Ministry of Business, Innovation & Employment COVID-19 Innovation Acceleration Fund, contract CIAF 0490.

## Supporting Information

Additional Supporting Information may be found in the online version of this article:

**Supplementary Fig. S1**. Neutralizing titer (NT50) values in the 140 CCP samples.

**Supplementary Fig. S2**. ROC curve of C19-kodecyte assay.

**Supplementary Table S1**. Characteristics of COVID-19 convalescent plasma donors.

**Supplementary Table S2**. Agreement and Cohenś kappa between C19-kodecyte and Ortho Total assays.

**Supplementary Table S3**. CCP donors with inconsistent results in the 5 assays.

### Web Resources

http://vassarstats.net/clin1.html#note

https://www.graphpad.com/quickcalcs/kappa2/

## References

1. WHO coronavirus disease (COVID-19) dashboard. Geneva: World Health Organization, 2020 (https://covid19.who.int/; accessed 5/27/2021).

2. Reuters. CanSino’s COVID-19 vaccine candidate approved for military use in China (https://www.reuters.com/article/us-health-coronavirus-china-vaccine/cansinos-covid-19-vaccine-candidate-approved-for-military-use-in-china-idUSKBN2400DZ; accessed 5/27/2021).

3. Ullah Z, Rahim Z. Putin says Russia has approved ‘world first’ Covid-19 vaccine. But questions over its safety remain (https://www.cnn.com/2020/08/11/europe/russia-coronavirus-vaccine-putin-intl/index.html; accessed 5/27/2021).

4. FDA. Pfizer-BioNTech COVID-19 vaccine (https://www.fda.gov/emergency-preparedness-and-response/coronavirus-disease-2019-covid-19/pfizer-biontech-covid-19-vaccine; accessed 5/27/2021).

5. Craven J. COVID-19 vaccine tracker (https://www.raps.org/news-and-articles/news-articles/2020/3/covid-19-vaccine-tracker; accessed 5/27/2021).

6. Denise M. Hinton, U.S. Food & Drug Admin., U.S. Dep’t of Health & Human Servs., Emergency Use Authorization for COVID-19 Convalescent Plasma (originally issued Aug. 23, 2020, and subsequently reissued with revisions) available at https://www.fda.gov/media/141477/download.

7. Rojas M, Rodríguez Y, Monsalve DM, et al. Convalescent plasma in Covid-19: possible mechanisms of action. Autoimmun Rev 2020; 19:102554.

8. Libster R, Pérez Marc G, Wappner D, et al. Early high-titer plasma therapy to prevent severe covid-19 in older adults. N Engl J Med 2021; 384:610–8.

9. Janiaud P, Axfors C, Schmitt AM, et al. Association of convalescent plasma treatment with clinical outcomes in patients with COVID-19: a systematic review and meta-analysis. Jama 2021; 325:1185–95.

10. Marks P. FDA In Brief: FDA updates Emergency Use Authorization for COVID-19 convalescent plasma to reflect new data (https://www.fda.gov/news-events/fda-brief/fda-brief-fda-updates-emergency-use-authorization-covid-19-convalescent-plasma-reflect-new-data; accessed 5/27/2021).

11. West R, Kobokovich A, Connell N, Gronvall GK. COVID-19 antibody tests: a valuable public health tool with limited relevance to individuals. Trends Microbiol 2021; 29:214–23.

12. FDA. Individual EUAs for serology tests for SARS-CoV-2 (https://www.fda.gov/medical-devices/coronavirus-disease-2019-covid-19-emergency-use-authorizations-medical-devices/in-vitro-diagnostics-euas-serology-and-other-adaptive-immune-response-tests-sars-cov-2; accessed 5/27/2021).

13. Nagappan R, Flegel WA, Srivastava K, et al. COVID-19 antibody screening with SARS-CoV-2 red cell kodecytes using routine serologic diagnostic platforms. Transfusion 2021; 61:1171–80.

14. De Giorgi V, West KA, Henning AN, et al. Anti-SARS-CoV-2 serology persistence over time in COVID-19 convalescent plasma donors. medRxiv 2021:2021.03.08.21253093.

15. Padoan A, Bonfante F, Pagliari M, et al. Analytical and clinical performances of five immunoassays for the detection of SARS-CoV-2 antibodies in comparison with neutralization activity. EBioMedicine 2020; 62:103101.

16. Walls AC, Park YJ, Tortorici MA, Wall A, McGuire AT, Veesler D. Structure, function, and antigenicity of the SARS-CoV-2 spike glycoprotein. Cell 2020; 181:281-92.e6.

17. Bennett RS, Postnikova EN, Liang J, et al. Scalable, micro-neutralization assay for qualitative assessment of SARS-CoV-2 (COVID-19) virus-neutralizing antibodies in human clinical samples. Viruses 2021; 13:893.

18. Marsh WL. Scoring of hemagglutination reactions. Transfusion 1972; 12:352–3.

19. Newcombe RG. Two-sided confidence intervals for the single proportion: comparison of seven methods. Stat Med 1998; 17:857–72.

20. Cohen J. A coefficient of agreement for nominal scales. Educ Psychol Meas 1960; 20:37–46.

21. Landis JR, Koch GG. The measurement of observer agreement for categorical data. Biometrics 1977; 33:159–74.

22. Ryzhov IM, Tuzikov AB, Nizovtsev AV. SARS-CoV-2 peptide bioconjugates designed for antibody diagnostics (in press). Bioconjugate Chemistry 2021.

23. Prus K, Alquist CR, Cancelas JA, Oh D. Decrease in serum antibodies to SARS-CoV-2 in convalescent plasma donors over time. Transfusion 2021; 61:651–4.

24. Yamayoshi S, Yasuhara A, Ito M, et al. Antibody titers against SARS-CoV-2 decline, but do not disappear for several months. EClinicalMedicine 2021; 32:100734.

25. Wajnberg A, Amanat F, Firpo A, et al. Robust neutralizing antibodies to SARS-CoV-2 infection persist for months. Science 2020; 370:1227–30.

26. Perreault J, Tremblay T, Fournier MJ, et al. Waning of SARS-CoV-2 RBD antibodies in longitudinal convalescent plasma samples within 4 months after symptom onset. Blood 2020; 136:2588–91.

27. Ibarrondo FJ, Fulcher JA, Goodman-Meza D, et al. Rapid decay of anti-SARS-CoV-2 antibodies in persons with mild Covid-19. N Engl J Med 2020; 383:1085–7.

28. van der Heide V. Neutralizing antibody response in mild COVID-19. Nat Rev Immunol 2020; 20:352.

29. Payne DC, Smith-Jeffcoat SE, Nowak G, et al. SARS-CoV-2 infections and serologic responses from a sample of U.S. navy service members - USS Theodore Roosevelt, April 2020. MMWR Morb Mortal Wkly Rep 2020; 69:714–21.

30. Kirkcaldy RD, King BA, Brooks JT. COVID-19 and postinfection immunity: limited evidence, many remaining questions. Jama 2020; 323:2245–6.

31. Wu F, Wang A, Liu M, et al. Neutralizing antibody responses to SARS-CoV-2 in a COVID-19 recovered patient cohort and their implications. medRxiv 2020:2020.03.30.20047365.

32. FDA. Template for test developers of serology tests that detect or correlate to neutralizing antibodies (https://www.fda.gov/media/146746/download; accessed 5/27/2021).

33. Heathcote D, Carroll T, Wang JJ, et al. Novel antibody screening cells, MUT+Mur kodecytes, created by attaching peptides onto red blood cells. Transfusion 2010; 50:635–41.

