## Supplementary for "COVID-19 antibody detection and assay performance using red cell agglutination"

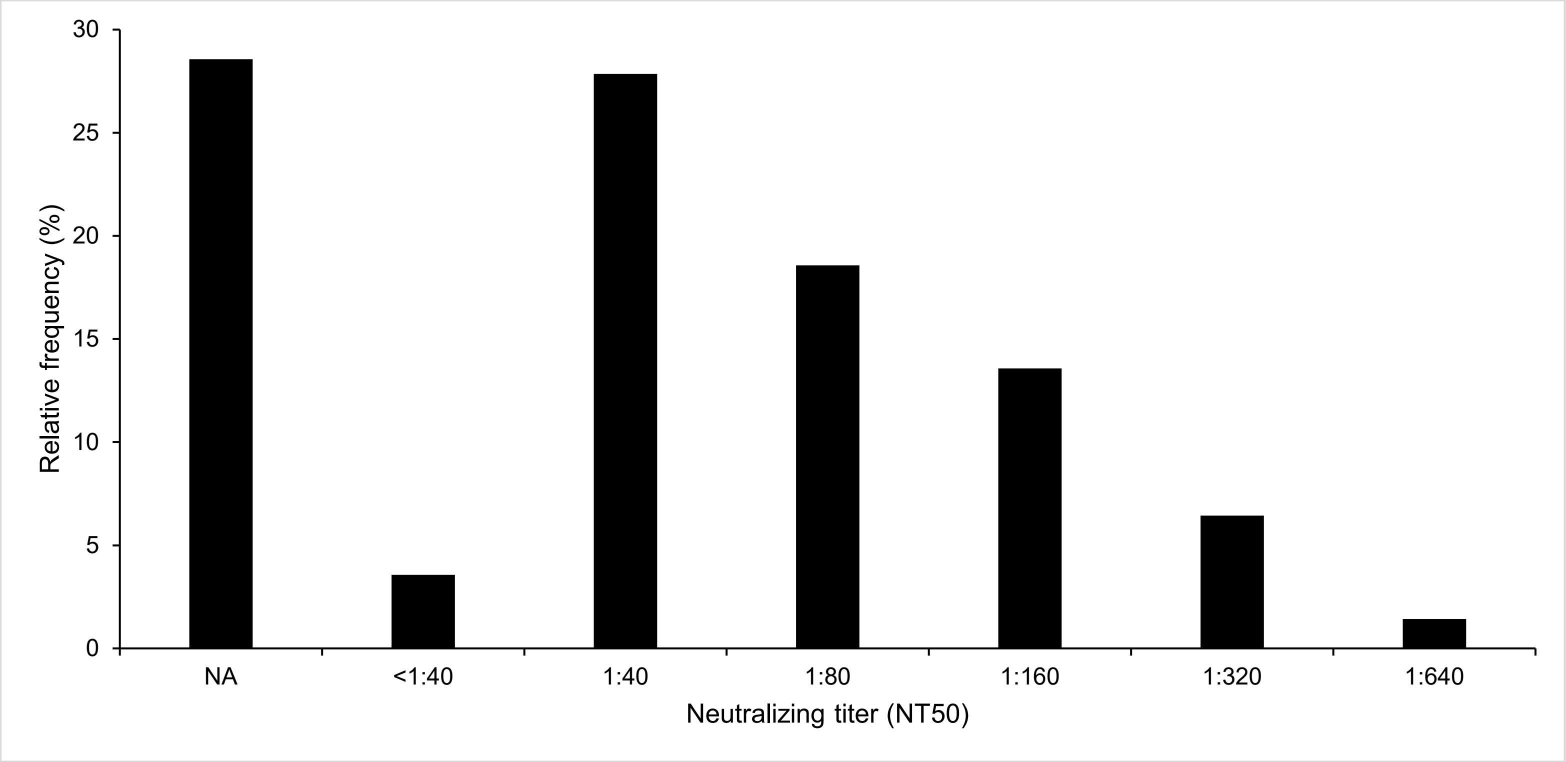
30
25
20
15
10
5
0
Relative frequency (%)
NA
<1:40
1:40
1:80
1:160
1:320
1:640
Neutralizing titer (NT50)

**Figure S1**

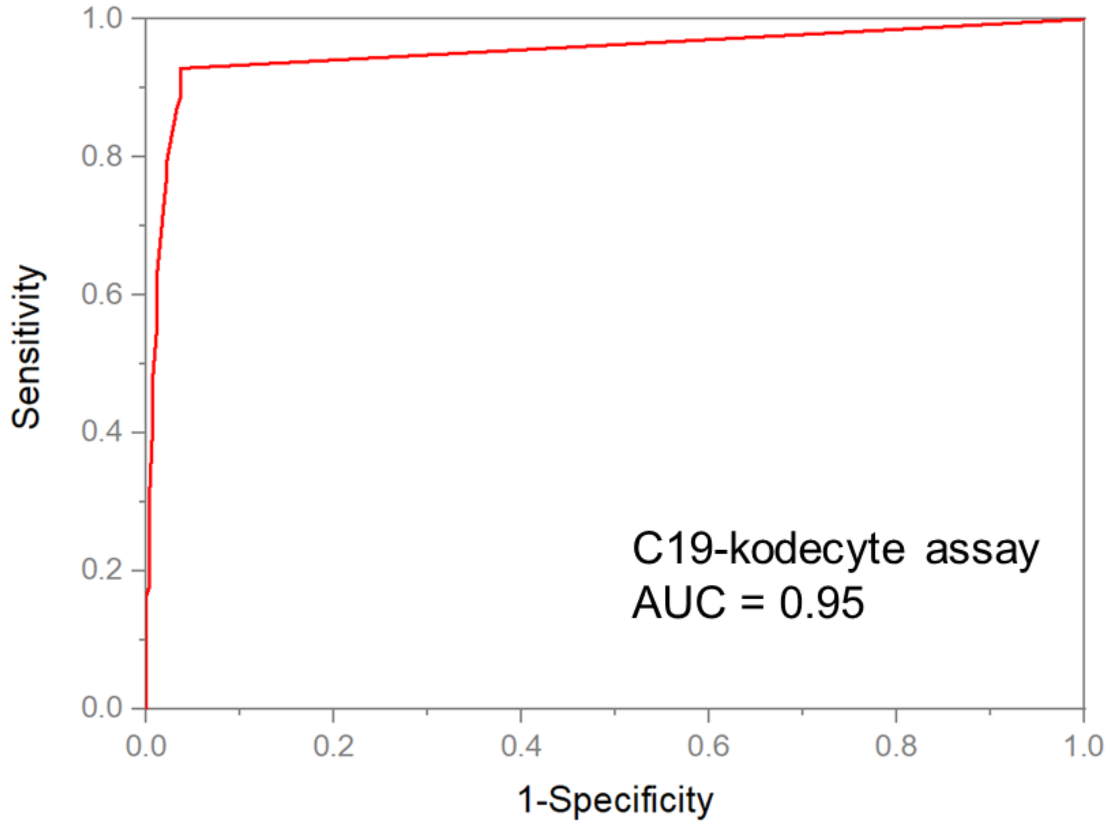
1.0
0.8
0.6
0.4
0.2
0.0
Sensitivity
0.0
0.2
0.4
0.6
0.8
1.0
1-Specificity
C19-kodecyte assay
AUC = 0.95

**Figure S2**

**Table S1.** Characteristics of COVID-19 convalescent plasma donors

| **Total number of convalescent plasma donors** | **n = 140** |
|---|---|
| Median time after symptom onset in days (range) | 94 (33 - 331) |
| Median time after PCR positive* testing in days (range) | 78 (3 - 292) |
| Convalescent plasma donors with | |
| No NT50 data %) | 40 (28.6) |
| NT50 <1:40 (%) | 5 (3.6) |
| NT50 >1:40 (%) | 95 (67.8) |
| **Total number of expected COVID-19 negative donors** | **n = 275** |
| Collected in 2020 (Ortho Total negative) | 125 |
| Collected in 2008 (no FDA authorized test, pre-COVID-19) | 150 |

* rRT-PCR for SARS-CoV-2 viral RNA

**Table S2.** C19-kodecyte and Ortho Total assay results: Agreement and Cohen´s kappa

| | C19-kodecyte assay | | |
|---|---|---|---|
| Ortho Total assay | Negative | Positive | Total (%) |
| Negative | 122 | 7 | 129 (48.7%) |
| Positive | 6 | 130 | 136 (51.3%) |
| Total (%) | 128 (48.3%) | 137 (51.7%) | 265 (100%)* |
| Agreement† | 95 % | | |

* rRT-PCR positive samples (140) + Ortho Total negative samples (125): 265
† Agreement: (122+130)/265 = 0.95

Cohen’s kappa coefficient (95% CI; standard error) = 0.90 (0.85-0.95; 0.027)
kappa coefficient <0.20 = poor agreement, 0.21 – 0.40 = fair agreement, 0.41 – 0.60 = moderate agreement, 0.61 – 0.80 = substantial agreement, and 0.81 – 1.00 = almost perfect agreement [21].

**Table S3.** CCP donors with inconsistent results among the 5 COVID-19 antibody assays

| | COVID-19 antibody assays | | | | | |
|---|---|---|---|---|---|---|
| Sample ID* | Ortho Total assay | Ortho IgG assay | Virus neutralizing assay | C19-kodecyte assay | Peptide 808-kodecyte assay | CCP donation after start of symptoms (days) |
| 1 | Positive | Positive | NA | Negative | Negative | 65 |
| 2 | Positive | Positive | Positive | Negative | Positive | 89 |
| 3 | Positive | Positive | NA | Negative | Negative | 85 |
| 4 | Positive | Positive | Positive | Negative | Positive | 77 |
| 5 | Positive | Positive | Positive | Negative | Positive | 88 |
| 6 | Positive | Positive | NA | Negative | Negative | 273 |
| 7 | Positive | Negative | Positive | Positive | Negative | NA† |
| 8 | Positive | Negative | NA | Positive | Negative | 52 |
| 9 | Positive | Negative | Positive | Positive | Positive | 65 |
| 10 | Positive | Negative | Positive | Positive | Negative | 47 |
| 11 | Positive | Negative | Positive | Positive | Positive | 97 |
| 12 | Positive | Negative | Positive | Positive | Negative | 110 |
| 13 | Positive | Negative | NA | Positive | Negative | 141 |
| 14 | Positive | Negative | NA | Negative | Negative | 33 |
| 15 | Negative | Negative | NA | Negative | Negative | 180 |
| 16 | Negative | Negative | NA | Negative | Negative | 90 |
| 17 | Negative | Negative | NA | Negative | Negative | 47 |

* Distinct donors (single donations)

† CCP donor never experienced symptoms (asymptomatic infection)

NA – not available
